# Benchmarking Artificial Intelligence vs General Practitioners Decision-Making in Same-Day Appointments Triage: A Mixed-Methods Study in UK Primary Care

**DOI:** 10.1101/2025.06.11.25329441

**Authors:** Sami Altalib, Eva Riboli-Sasco, Mahmoud Al Ammouri, Hannah Gibson, Katherine Leung, Annabelle Painter, Ragu Rajan, Austen El-Osta

## Abstract

**Background:** Artificial intelligence (AI) is increasingly used to support clinical decision-making, particularly in primary care triage. However, few studies have benchmarked AI triage tools against general practitioner (GP) assessments in real-world settings. This study evaluated the agreement between an AI-enabled triage tool (Visiba Triage) and GP urgency ratings for same-day appointment requests. Secondary aims included assessing perceptions of safety, accuracy and usability from both clinician and patient perspectives.

**Methods:** A mixed-methods study was conducted using data from patients requesting SDA between January and June 2024. Urgency scores generated by the Visiba Triage AI tool based on a modified Manchester Triage System were compared to GP-assigned ratings using Spearman’s rank correlation and Cohen’s kappa. Ordinal logistic regression assessed associations between demographics and patient satisfaction. Thematic analysis of interviews with eight GPs explored perceptions of the AI tool’s performance.

**Results:** A total of 649 participants were included in this study. The majority were females and of White ethnicity. There was a strong correlation between AI and GP urgency ratings (ρ=0.796, *p* 0.001), with 83.7% categorical agreement across eight urgency levels (κ 0.69, *p* 0.001). The AI system demonstrated safety-conscious design, with a greater likelihood of over-triage whilst rarely under-triaging. No cases deemed non-urgent by AI were later reclassified as emergencies by GPs. Qualitative findings supported the quantitative results, highlighting perceived accuracy and safety. Current limitations include suboptimal integration with patient medical records. Patient satisfaction varied significantly by age, with older adults (60+) reporting lower satisfaction (aOR 0.25, 95% CI 0.12-0.52).

**Conclusion:** This study demonstrates that AI-enabled triage can closely mirror clinical judgement in a primary care setting, offering a safe, scalable solution to manage demand for same-day care. Safe adoption of AI triage tools in healthcare should include real-world assessment and benchmarking against consensus clinician judgement in real-time.

**Key Takeaways:**

- AI-enabled triage tools can achieve substantial agreement with GP urgency assessments, with 84% categorical concordance and no observed cases of significant under-triage.
- The AI model demonstrated a safety-conscious design, favouring over-triage to reduce patient safety risks, especially in emergency scenarios.
- Older adults reported significantly lower satisfaction with AI triage, highlighting the need to address digital literacy and inclusion when implementing such tools.
- GPs expressed high confidence in AI performance at acuity extremes, particularly for self-care and emergency cases, though noted contextual limitations without EHR integration.
- This real-world study highlights the potential of AI triage to enhance clinical efficiency, particularly in managing same-day appointment demand in overstretched systems like the NHS.
- Ongoing clinician oversight remains essential to mitigate AI limitations in complex cases and ensure equitable, safe deployment at scale.

## Introduction

The front door of the NHS conducts 1.54 million patient contacts per day (1,2). An estimated 60% of this activity across Primary Care and Integrated Urgent Care is from patients requesting same-day services (3). Current practice does not routinely screen the appropriateness of same-day activity before allocation.

In recent years, artificial intelligence (AI)-enabled triage tools have emerged which claim to be able to determine where patients have a same-day need or not. These systems, typically integrated into patient-facing apps or online forms, guide users through structured symptom checkers and generate urgency recommendations. The COVID-19 pandemic catalysed the rapid adoption of “total triage” and “digital-first” models in primary care, many of which now blend AI tools with traditional workflows (4). The hope is that AI triage can maintain safety and accuracy while improving access and efficiency in primary care (5).

AI-assisted triage and decision support tools are already adopted at scale in countries such as Sweden and Finland to address the growing demands on healthcare systems, improve the accuracy of service user assessments and optimise resource allocation (6). The most advanced tools used for this purpose are medically trained probabilistic graphical models (PGMs) that assist general practitioners (GPs) or other clinicians in making more informed decisions about triage urgency to enhance service user outcomes and operational efficiency. One of the key advantages of using a medically trained PGM is the capability to replicate a clinician’s quality of assessment, quickly and efficiently. These systems can take a patient history, identify symptom patterns and understand disease correlations akin to what a clinician is trained to do (7). PGMs are designed to be dynamic and adaptable, allowing them to be updated with new data, medical knowledge and clinical guidelines. This ongoing refinement ensures that the system’s recommendations remain explainable, relevant, evidence-based and aligned with current best practices. In primary care, where conditions are often non-specific and symptoms can be vague, the ability of AI to provide tailored, evidence-based recommendations is particularly valuable (8).

Earlier studies found that 15-30% of Emergency Department attendances in the NHS were amenable to being safely managed in primary care (9) and that three-quarters of NHS patients did not know which service to present to at first contact (10). The ability for the PGM to replicate a clinician’s quality of assessment and support intelligent navigation to an appropriate level of care is potentially revolutionary for health systems where poor service user navigation and workforce shortages are systemic. Despite the opportunity, the implementation of AI models as triage and navigation tools has several challenges including concerns about liability, triage accuracy, algorithmic bias and the transparency of AI decision-making processes. The integration of AI into clinical workflows also requires careful planning to ensure that these tools complement rather than replace the expertise of healthcare professionals (7).

### AI-enabled triage tool primary care pilot

This study looked at the deployment of a medically trained PGM in an English NHS primary care setting. The AI-enabled triage tool is a medically trained AI model based on a probabilistic graphical network. It is designed to be continuously updated by clinicians based on real-world feedback data and updates to clinical practice, ensuring the model reflects evolving medical knowledge. Despite these advances, there remains a paucity of real-world evaluations comparing AI-generated triage decisions to real-time clinical assessments under operational NHS conditions, which is considered to be the gold standard of safety (11).

The Visiba Triage AI-enabled tool is already deployed at scale in Nordic health systems. Importantly, its integration into clinical workflows allows for clinical oversight and contextualisation of AI recommendations mitigating safety risks. Given the rising importance of PGMs, there is a need to evaluate the congruence of effective AI Triage tools in supporting patients requesting same-day appointments in the UK.

The aim of this mixed methods study was to evaluate the concordance between the AI-enabled triage tool and GP-assigned urgency ratings for same-day appointment requests in UK primary care. We also explored GP perceptions of safety and performance and analysed patient feedback to assess demographic patterns in user satisfaction. The overarching objective was to determine whether AI-enabled triage can safely and acceptably replicate clinical judgment at scale.

## Methods

### Study design

We conducted an explanatory sequential mixed-methods study comprising a cross-sectional quantitative analysis followed by qualitative interviews. The design was chosen to assess both the concordance between AI and GP triage decisions and to contextualise these findings through clinician insights.

### Setting and intervention

The study was conducted across a partnership of four English NHS practices in East Sussex between January and June 2024. Prior to implementation, patients requesting same-day appointments (SDA) were triaged manually by reception staff and allocated to duty doctors until all slots were filled. Patients who called after this cut-off were redirected to NHS 111, advised to try again the next day or else added as an overflow patient to the GP’s worklist at the discretion of the receptionist.

During the pilot, the following hybrid model was introduced (**Figure 1**): patients requesting SDAs after the cut-off time received an SMS link to complete an AI-enabled triage. Those unable to access the system due to digital or linguistic barriers were supported by reception staff acting as proxies. Once the information was collected, the AI system generated an urgency score based on a modified Manchester Triage System (MMTS) which categorised patients into one of eight urgency levels based on the maximum acceptable waiting time before consultation (**Supplementary Table 1 in Appendix).** Scores 1 to 3 denote emergencies, where patients must be seen within 0, 10 or 60 minutes, respectively. These categories reflect life-threatening or time-critical conditions requiring immediate or near-immediate intervention. Scores 4 and 5 fall under the ’within 24 hours’ category, with waiting times of 4 hours and 1 day, suitable for less acute but still clinically significant conditions. Scores 6 and 7 represent ’non-urgent’ cases with recommended waiting periods of 1 week and 1 month, respectively, where the clinical concern is minor or stable. Finally, score 8 was designated for ’self-care’, indicating that the patient can manage their condition independently without professional intervention. This stratified system supports efficient resource allocation and prioritisation of care based on clinical urgency. The urgency score was then presented and reviewed by a GP with access to the patient’s electronic health record (EHR) to support with clinician-led decision making. After considering contextual factors and reviewing the EHR of the patient, a GP would decide on the final urgency score to inform next steps. The GP also submitted feedback on whether they agreed with the AI system’s urgency score and if they disagreed, what urgency score they felt was more clinically appropriate. The patient feedback score was also collected and analysed.

**Figure 1:**
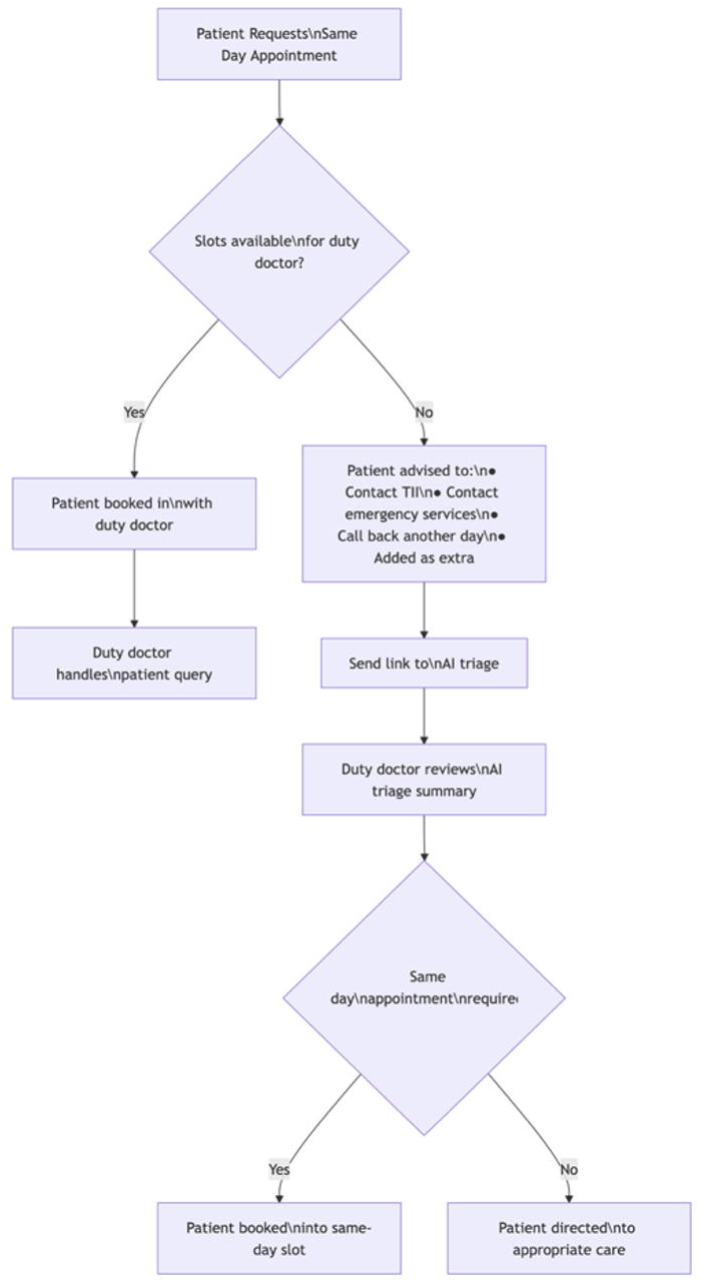
Flowchart representation of the approach to managing same-day appointment requests before and after introduction of the AI-Triage tool.

### Participants

For the quantitative phase, anonymised secondary data collected by the AI-enabled triage tool was obtained from participating sites. The dataset comprised 2,671 rows of anonymised patients who used the AI triage tool to request SDA during the 6-month study period. No exclusions were applied based on age, gender or ethnicity.

For the qualitative arm, a convenience sample of eight GPs who had used the system for 6-8 months between 1 January and 30 June 2024 were recruited for semi-structured interviews. All participants were practising within the study site and provided written informed consent.

### Sampling and sample size

For the quantitative arm, simple random sampling was used to select users for GP triage review. A minimum sample size of 532 was calculated using OpenEpi (99% CI, 5% precision, population size=2,671, anticipated frequency=50%). This ensured robust power for correlation and regression analyses.

### Data collection

Demographic data included patient age, gender, ethnicity and whether the participant had a carer were retrieved from the EHR of patients using the AI-enabled triage tool during the 6-month period (Jan-Jun 2024). For each patient, data including the AI triage tool’s score and the GP triage score based on the MMTS were collected.

For the qualitative arm, a convenience sample of six GPs (4 males, 2 females) were interviewed after being approached by the research team with an ethically approved participant information sheet outlining the research project’s objectives. Following consent, semi-structured online interviews and focus group discussions were conducted on 12 and 13 August 2024 via MS Teams. The interviews were audio recorded and transcribed ad verbatim. Respondents were assured of their voluntary participation and their right to withdraw at any point. All participants had been using the AI triage tool for a range of 6-8 months. Interviews were led by AEO and ERS. Interviews lasted between 45 to 55 minutes and were guided by a semi-structured interview guide which was reviewed internally by the research team **(Supplementary Table 2 in Appendix).** No repeat interviews were carried out. Only two researchers (AEO and ERS) had access to the transcripts which were stored on password-protected Imperial College London’s secure online environment. Transcripts were not returned to participants, and no feedback was requested from them regarding the analysis.

### Data analysis

For the quantitative arm, descriptive statistics were used to summarise findings in frequencies and percentages. The primary analysis involved comparing the urgency ratings assigned by the AI model and with those provided by GPs. A Spearman’s rank correlation analysis was conducted to assess the strength of the relationship between the two sets of urgency ratings. The correlation coefficient (ρ) and p-value were calculated to determine the statistical significance of the observed correlations. A scatter plot was used to visualise the relationship between triage scores assigned by AI and GPs. To mitigate overplotting and enhance data point visibility, we applied jittering to both axes. This technique involves adding small random noise to data points, effectively spreading out overlapping points and providing a clearer representation of data density and distribution patterns. Then, the scores by the AI and GPs were categorised. The categories were defined as follows: emergency (scores 1-3), within 24 hours (scores 4-5) and non-urgent (scores 6-8) (See Table 1). Overtriage was defined as when the GP’s urgency score was one category lower than the AI’s urgency score and significant overtriage as when the GP’s urgency score was two categories lower than the AI’s urgency score. Similarly, undertriage was defined as when the GP’s urgency score was one category higher than the AI’s urgency score and significant undertriage as when the GP’s urgency score was two categories higher than the AI’s urgency score.

**Table 1:** Demographic characteristics of patients who requested Same-Day Appointment.

| Variable (N=649) | Frequency | Percentage |
| --- | --- | --- |
| **Age** |  |  |
| 0-9 | 94 | 14.6 |
| 10-19 | 57 | 8.89 |
| 20-39 | 113 | 17.6 |
| 40-59 | 148 | 23.1 |
| 60-79 | 155 | 24.1 |
| 80 & above | 75 | 11.7 |
| Missing | 7 | - |
| **Gender** |  |  |
| Male | 218 | 34.0 |
| Female | 424 | 66.0 |
| Missing | 7 | - |
| **Ethnicity** |  |  |
| White | 498 | 98.8 |
| Asian / Asian British | 3 | 0.6 |
| Mixed | 3 | 0.6 |
| Missing | 145 | - |
| **Has Carer** |  |  |
| Yes | 6 | 0.9 |
| No | 636 | 99.1 |
| Missing | 369 | - |

Cohen’s Kappa was used to measure the agreement between the two ratings. Unadjusted and adjusted ordinal logistic regression models were used to test the association between demographics and users’ feedback scores. All analyses were performed using STATA, version 17 (StataCorp LP, College Station, TX, USA).

For the qualitative interview component, transcripts were analysed thematically following the Framework Method (12, 13). Emergent themes from the contextual data were classified around the concepts of perceived accuracy, safety and acceptability.

The quantitative component of the study was reported in accordance with the Strengthening the Reporting of Observational Studies in Epidemiology (STROBE) checklist. For the qualitative component, we followed the Consolidated Criteria for Reporting Qualitative Research (COREQ) 32-item checklist.

### Ethical considerations

The study received ethics approval from Imperial College Research Ethics Committee (ICREC #7106912). Participants consented to take part before the start of interviews. Participants were free to withdraw from the interview at any time. Interview data was pseudonymised. The interviews were transcribed with the principle of anonymity in mind and transcriptions were not outsourced, therefore no confidentiality agreements were required. All data generated or analysed during this study are included in this published article.

## Results

### Participant characteristics

A total of 649 participants were included in the analysis. **Table 1** presents full demographic characteristics of the cohort. The majority were female (66.0%) and most identified as White (98.8%). Only 0.9% of patients indicated they had a carer. The most represented age group was 60-79 years (24.1%), followed by 40-59 years (23.1%) and 20-39 years (17.6%). A substantial proportion of triage requests for those aged under 19 years were assumed to be submitted by parents or guardians.

### Comparison of AI and GP triage scores

Urgency ratings from the AI-enabled triage tool and GPs were compared across the 8- point MMTS. **Table 2** shows the distribution of urgency ratings by both methods. The AI system assigned most cases to level 5 (1-day review, 36.4%) or level 4 (4-hour review, 20.5%). GPs similarly assigned most patients to level 5 (46.4%) or level 4 (21.9%). Notably, the triage tool classified 10.5% of requests as requiring immediate attention (level 1), compared to 5.7% by GPs.

**Table 2:** Distribution of Triage Urgency Scores Assigned by the AI-Enabled Triage Tool and General Practitioners to Patients Requesting a Same-Day Appointment.

| Variable (N= 649) | Frequency | Percentage |
| --- | --- | --- |
| **AI Triage Urgency** |  |  |
| 1 (0 min) | 68 | 10.5 |
| 2 (10 min) | 4 | 0.6 |
| 3 (60 min) | 6 | 0.9 |
| 4 (4 hours) | 133 | 20.5 |
| 5 (1 day) | 236 | 36.4 |
| 6 (1 week) | 118 | 18.2 |
| 7 (1 month) | 9 | 1.4 |
| 8 (Self-care) | 75 | 11.6 |
| **GP Feedback Urgency** |  |  |
| 1 (0 min) | 37 | 5.7 |
| 2 (10 min) | 2 | 0.3 |
| 3 (60 min) | 7 | 1.1 |
| 4 (4 hours) | 142 | 21.9 |
| 5 (1 day) | 301 | 46.4 |
| 6 (1 week) | 100 | 15.4 |
| 7 (1 month) | 4 | 0.6 |
| 8 (Self-care) | 56 | 8.6 |

### Agreement between AI and GP triage decisions

There was a strong positive correlation between the tool and GP-assigned triage scores (Spearman’s ρ=0.796, *p*<0.001). **Figure 2** visualises the score alignment. Categorising the scores into clinical urgency levels (emergency (1–3), urgent within 24 hours (4–5) and non-urgent (6–8)) revealed that 83.7% of cases were classified identically (**Table 3**).

**Figure 2:**
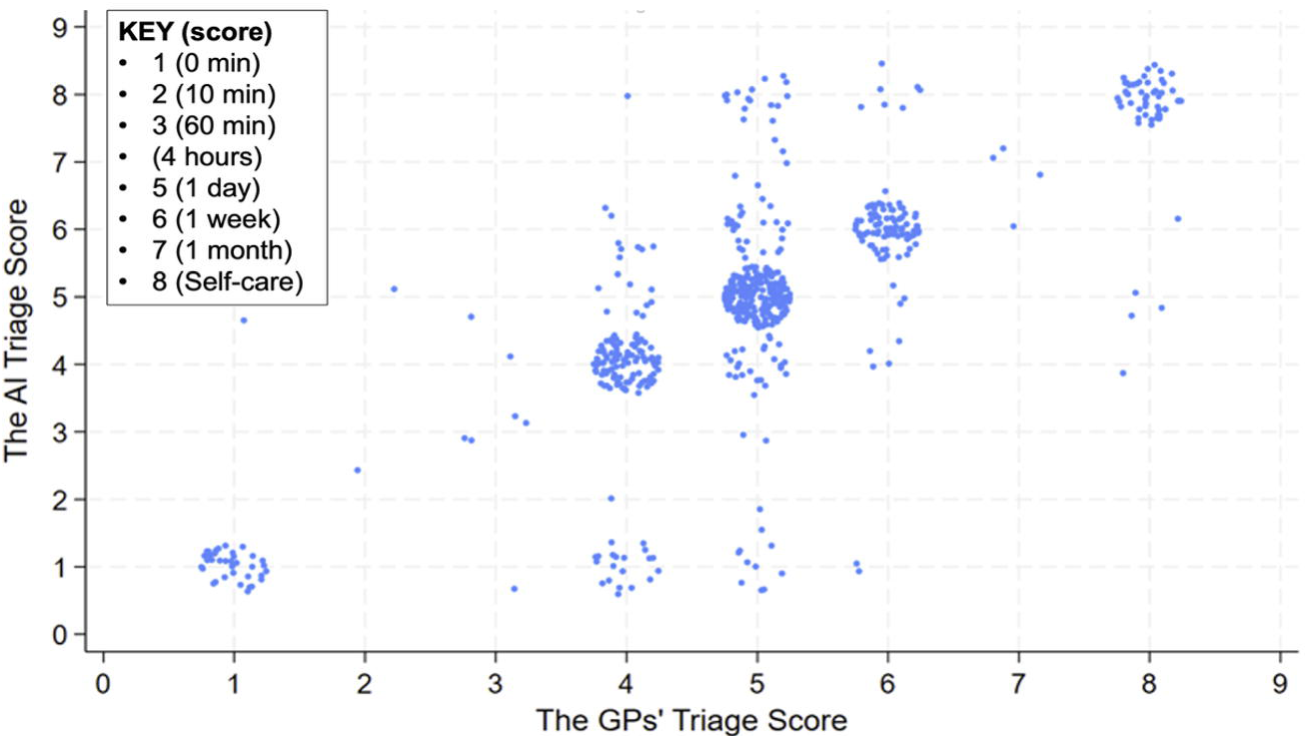
Correlation Between AI and GP Triage Urgency Scores for Same-Day Appointment Requests.

**Table 3:** Distribution of triage urgency category agreement between AI and GP for Same-Day Appointment requests.

| Triage urgency category | Frequency | Percentage | Confidence intervals |
| --- | --- | --- | --- |
| Significant undertriage | 0 | 0.0% | (0.0% - 0.6%) |
| Undertriage | 59 | 9.1% | (7.1% - 11.5%) |
| Correct triage | 543 | 83.7% | (80.6% - 86.3%) |
| Overtriage | 45 | 6.9% | (5.2% - 9.2%) |
| Significant overtriage | 2 | 0.3% | (0.1% - 1.1%) |
| **Total** | **649** | **100%** | - |

The Cohen’s κ was 0.69 (SE=0.03, p<0.001), indicating substantial agreement beyond chance. According to Landis and Koch (1977), kappa values between 0.61 and 0.80 denote “substantial” agreement, suggesting that the AI system closely approximates GP assessments within this triage context (14).

As shown in **Figure 2**, the triage urgency scores generated by the AI system and GPs were closely aligned, indicating consistent triage levels between the two. That the dispersion remained minimal reflected strong agreement, even across varying urgency categories. Out of all cases submitted for same-day appointment requests, there was no significant undertriage. Only 9.1% of all cases were undertriaged but none were associated with adverse safety events during the study period. The observed cases of undertriage primarily involved presentations where clinical context such as comorbidities, frailty or recent deterioration could not be fully appreciated by the AI due to inability to extract data from the EHR directly.

**Table 4** shows the confusion matrix of urgency assessments. From this matrix, we can calculate specific triage category sensitivity by normalising by GP triage assessment. The AI triage tool correctly detected patients who required (i) emergency care in 91.3% of cases, same-day care (within 24 hours) in 79.9% of cases and (iii) patients who required non-urgent care in 91.9% of cases.

**Table 4:**
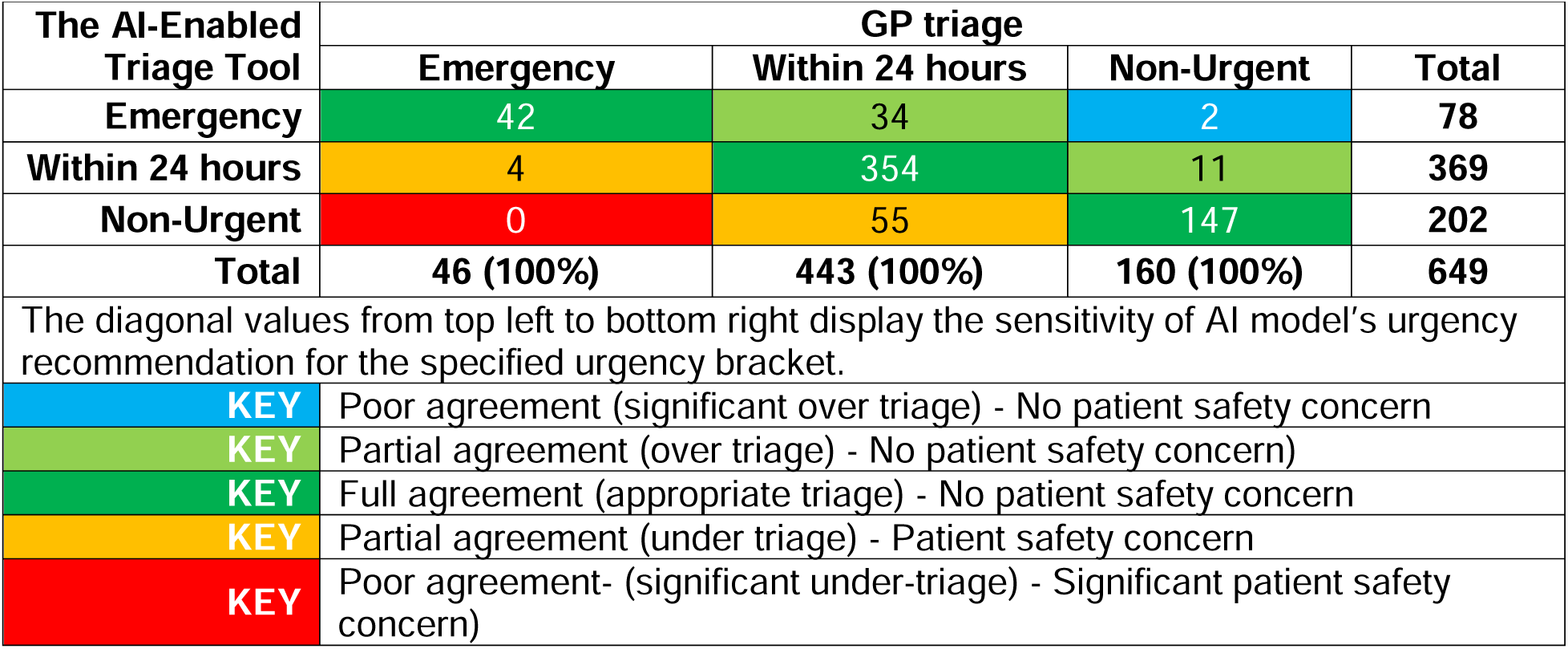
Confusion matrix of urgency assessments.

### GP perceptions of AI triage accuracy and safety

Interviews with eight GPs confirmed that the AI Triage tool was generally accurate and safe, with all respondents reporting high confidence in AI triage decisions for non-urgent and emergency presentations.

### Inherent safety netting within of AI triage

GPs consistently described the decision support tool as "appropriately conservative" and generally safe, especially in cases requiring urgent attention. Most felt reassured by the system’s cautious approach.

*“I think we’re all quite reassured that it was conservative rather than anything else really… very rarely I would say that something was actually more urgent than the software had suggested.”* (GP2, M)

Most GPs thought that *“the extremes* - i.e. - *either the ones that are high priority or self-care that sometimes I think it’s slightly off, but it’s the ones in the middle that generally perform relatively well”* (GP1, M), whereas some felt that *“the extremes are fine. It’s in the between that can differ based on the history that’s provided”* (GP5, M).

### Limitations in contextual awareness

Despite overall trust in the AI tool, clinicians noted that its inability to extract data from the EHR reduced its effectiveness in complex cases. Age, comorbidities and risk stratification were seen as critical gaps.

*“We need to have integration with the patients past medical history for it to be more effective. You know, a cough in an 88-year-old smoker is very different to a cough in a 21-year-old non-smoker who’s fit and well. So that integration and that sort of risk stratification of the patient’s past medical history is I think an essential part of any platform going forward”.* (GP6, M)

This limitation was mitigated to some extent because the standard operating procedure required that all AI Triage recommendations are revised by a GP who has sight of the patient’s EHR. This approach was adopted to inform decision making regarding the triage urgency as patient level data from EHR was not available to the AI decision support tool.

***“****… because [the AI Triage tool doesn’t have the context and information from the clinical system, there may be an elderly frail person for whom it may say self-care, but you know that they’re quite vulnerable and you may then upgrade them to an appointment but not maybe on that day but later on in that week. So, it’s in those contexts where we kind of change the triage”.* (GP1, M)

### Equity, digital literacy and proxy use

GPs highlighted the challenges posed by proxy submissions (e.g. parents submitting on behalf of children), non-native English speakers and older adults with limited digital literacy.

*“The fact that not all the time the patients describe things in detail. There can be requests which are literally one or two words which makes it rather inconvenient. So, we just kind of go back to them and it’s more of a learning process for the patient as well to kind of make sure that they use the opportunity of using the link to provide us with the maximum information.”* (GP7, M)

### Patient acceptability and satisfaction

Of the 649 patients, only 280 (43.1%) submitted feedback on their AI triage experience **(Supplementary Table 3 in Appendix)**. Among those, 65% rated the experience as “very good” or “excellent” (score 4 or 5). However, older adults reported lower satisfaction. However, as only 43.1% submitted feedback, this raises the potential for response bias. We therefore acknowledge that patients who provided feedback may represent individuals with either particularly positive or negative experiences, thus limiting the generalisability of satisfaction findings. Additionally, no comparative demographic analysis was performed between respondents and non-respondents, which limits the interpretation of observed satisfaction trends.

**Table 5** presents the regression analysis of demographic predictors of feedback. Compared to the 20-39 year reference group, those aged 60-79 had significantly lower odds of giving a high rating (aOR 0.25, 95% CI 0.12-0.52), as did those aged 80+ (aOR 0.14, 95% CI 0.05-0.39). Satisfaction did not differ by gender.

**Table 5:**
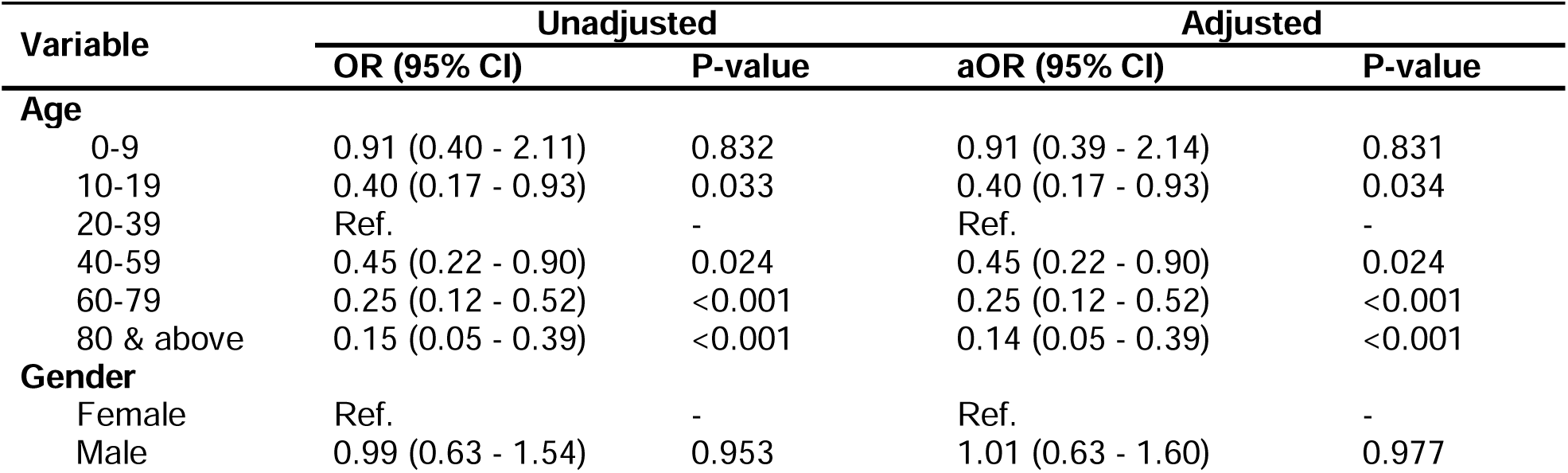
Ordinal Logistic Regression of Patient Demographics and Satisfaction With the AI-Enabled Triage Tool.

The 20-39-year age group was selected as the reference category due to its relatively high satisfaction ratings and larger representation in the sample. Female gender was chosen as the reference due to its predominance in the study population (66%), allowing for more stable estimates.

### GP reflections on patient experience

GPs reported that younger adults and parents were most receptive to the system. In contrast, older patients often needed support often due to lower digital literacy.

GPs reported that mothers with young children appeared to be particularly receptive and enthusiastic about AI Triage, since this additional (rather than a mandatory) step maintained the possibility for patients to book appointments in the traditional via phone or in person. GPs also reported that service user hesitance tended to dissipate as they became accustomed to using the system and could see the benefits of using it.

***“****I think generally older people find it more of a struggle. Initially especially, I think there’s probably a little bit more resistance by older patients who were less Internet savvy. But I think we’d have a lot more resistance if that was the only way we could book appointments. The fact that they can still phone up or they can come in and they can make an appointment” **GP4, F***

*“I think this is [why] we’re only using [the AI-enabled triage tool] for those on the day, you know, things that people feel are urgent for that day and I think that patients, I don’t know that they appreciate it but having spoken to other practises, I think we provide quite a good service through this”*.

*“I hear a lot of feedback from other surgeries, who go through a system like this for every single appointment that’s booked and I think personally that would be quite challenging [and] would hit a lot of resistance.”*

## Discussion

### Summary of main findings

This mixed-methods study provides robust evidence for incorporating the utility of an AI- enabled triage tool to support the safe and effective management of same-day appointment requests within primary care. Quantitative analysis revealed a high degree of concordance between urgency scores assigned by the AI tool and those determined by GPs, with a Spearman’s rank correlation coefficient of 0.796 (p<0.001) and a Cohen’s κ of 0.69 (p<0.001) indicating substantial agreement. These findings highlight the potential of AI systems to approximate clinical decision-making in real-world operational contexts and to support the timely allocation of resources in overstretched healthcare environments.

The system demonstrated particularly strong performance in triaging non-urgent cases, with alignment between AI and GP assessments observed in 92% of such instances. Importantly, no cases designated by the AI as non-urgent were subsequently reclassified by clinicians as emergencies, indicating a high negative predictive value and a safety-first algorithmic design. Nevertheless, the AI occasionally undertriaged non-emergency patients, particularly older adults and those with complex medical needs. This was largely attributed to the tool’s current inability to access the EHR and use contextual data such as the presence of multimorbidity, frailty and other important medical history.

The qualitative component of the study supported these quantitative findings with all GPs expressing broad confidence in the accuracy and safety of the AI triage tool particularly at the extremes of acuity (i.e., self-care and emergency categories). The system’s conservative bias, manifesting in a tendency to over-triage, was widely regarded as a safety-enhancing feature rather than a liability, with clinicians valuing this as a safeguard in ambiguous or borderline presentations.

In terms of user experience, the AI system was generally well-received: among those who submitted feedback, 65% rated their interaction as “very good” or “excellent.” Notably, satisfaction varied significantly by age, with older adults (60+) reporting markedly lower satisfaction scores, highlighting a persistent digital divide in healthcare engagement. GPs corroborated these findings, noting that younger adults and parents of young children were more comfortable with the technology, whereas older or digitally excluded populations often required additional support or defaulted to alternative booking methods.

In summary, the AI-enabled triage tool exhibited high reliability in mirroring clinician urgency assessments, offering a scalable and safe adjunct to existing triage pathways in primary care. Its strength lies in its conservative design and alignment with GP decision-making, particularly in non-urgent scenarios. However, its current limitations include lack of EHR data extraction, contextual blind spots and digital access disparities. These findings affirm the importance of maintaining clinician oversight and emphasise the need for iterative refinement to optimise AI systems for equitable, context-aware and patient-centred deployment in real-world care settings.

### Comparison to existing literature

The integration of AI into healthcare, particularly in triage systems, is a rapidly growing area of research and practice (5, 6, 15). The findings from this study align with a growing body of literature that highlights the potential for AI to enhance efficiency, reduce clinician workload and improve the timeliness and accuracy of patient management, particularly in urgent and primary care settings.

Unlike studies that rely on clinical vignettes or synthetic datasets, this evaluation used real-world patient data and was benchmarked against consensus clinical judgement in live practice. While vignette-based assessments offer control and repeatability, they lack the complexity and contextual variability of real-world encounters. Clinical judgment incorporates nuanced information such as patient history, comorbidities, communication subtleties and clinical intuition factors that are often absent from artificial test scenarios. By using GP triage ratings as the reference standard in an operational setting, this study adopts an evaluation approach increasingly regarded as best practice in AI research, as it mirrors how the technology would be used in real-world workflows and emphasises patient safety, clinician trust and system usability (16).

### Performance relative to symptom checkers

The AI-enabled triage tool demonstrated robust triage performance, with substantial agreement with GP assessments. Its performance is similar to or surpasses that of several commercially available symptom checkers assessed in prior studies. In particular, the tool’s categorical sensitivity for emergency and 24-hour care far surpasses that reported in Hill et al.’s review of 19 symptom checkers (63% and 56%, respectively). The tool’s categorical triage agreement of 84% falls in the upper end of 49-90% which is the triage accuracy from Wallace et al. 2024’s systematic review of digital symptom checker tools (17).

### Contribution to the field

Overall, this study contributes critical evidence to the practical deployment of AI triage tools in real-world primary care. In contrast to the controlled or retrospective nature of many prior studies, our findings provide a pragmatic evaluation of AI performance during live triage scenarios, highlighting both opportunities and constraints. The alignment with clinical judgement, the risk-averse nature of the AI and the system’s acceptability by clinicians position it as a promising complement to existing triage workflows.

### Implications for research, policy and practice

The findings of this study suggest several avenues for future research. Further research is needed to examine the long-term impact of AI-enabled triage systems on service user outcomes, healthcare efficiency and GP workload. This could involve longitudinal studies that track the use of AI triage over time, as well as randomized controlled trials that compare outcomes between AI-assisted and traditional triage methods.

Policymakers should consider the integration of AI-enabled triage tools into national healthcare frameworks, particularly in systems like the NHS that face significant challenges in managing service user flow and resource allocation. The demonstrated ability for an AI-enabled triage tool to strongly and safely align with a GP’s assessment, presents the opportunity for intelligent navigation within health systems. The system-wide benefits of intelligent navigation were explored in detail by The Tony Blair Institute, with an estimated saving to the UK health system of £340m a year (10). The ability for the AI model to continually collect performance data in real-time is considered a critical safety feature and the gold standard of AI-enabled triage performance monitoring.

For healthcare providers, the integration of AI-enabled triage tools represents an opportunity to enhance clinical decision-making and improve service user care. The findings of this study suggest that these systems can reliably support GPs in identifying and prioritising urgent cases, potentially reducing wait times and improving service user outcomes. Providers must also remain vigilant in monitoring AI system performance and should be prepared to intervene when discrepancies arise.

### Strengths and limitations

A key strength of this study is its use of a large, real-world dataset drawn from routine primary care activity, which enhances the external validity and translational relevance of the findings. The mixed-methods design combining quantitative analyses with qualitative insights from experienced GPs allowed for a holistic appraisal of both technical accuracy and user acceptability. Importantly, the evaluation was situated in a live clinical workflow, rather than a simulated or retrospective environment, thereby capturing the practical realities of AI deployment in routine NHS care. The high observed agreement between AI and GP urgency assessments (Cohen’s κ=0.69) further strengthens the evidence base for AI-enabled triage as a reliable decision-support tool in same-day appointment management.

The principal limitation of this study was that the evaluation was conducted at an individual NHS site, limiting generalisability to other regions, populations or care models. Replicating this analysis across diverse healthcare settings, urban vs rural, digitally engaged vs digitally excluded populations and across varying socioeconomic contexts will be essential to determine the broader applicability of these findings.

Second is the issue of data quality and completeness. Missing values for key demographic variables such as ethnicity, as well as the low response rate of user feedback limits the generalisability of some findings. While no imputation was performed, future evaluations should ensure complete demographic data collection to enable more nuanced analysis of equity and inclusion. The extent of missingness may also introduce selection bias, particularly if certain groups were less likely to complete digital triage forms fully.

Third, although no emergency cases were under-triaged, the study did not systematically assess or follow up on potential harms associated with delayed or inappropriate triage. Furthermore, the retrospective design precludes causal inference and reliance on secondary data may introduce unmeasured confounding variables, such as symptom severity or patient communication style, which could influence both AI and clinician decision-making.

Fourth, the study only examined users seeking same-day care, excluding those pursuing routine, preventative, or chronic disease management. This limits the findings to a subset of primary care interactions and may not reflect the performance of the AI system in broader clinical scenarios. Moreover, only 43% of patients submitted feedback, introducing potential response bias. Those most or least satisfied may have been disproportionately represented and no comparative analysis was conducted between respondents and non-respondents.

Fifth, the use of proxy responders (e.g., parents or carers completing triage on behalf of patients) introduces a risk of misclassification or misrepresentation of symptoms.

Finally, the study did not include qualitative data from patients, which limits the depth of insight into user experience. Patient voices through interviews or focus groups would provide a more comprehensive understanding of acceptability, usability and perceived barriers to engagement.

## Conclusion

This study provides compelling evidence that a medically trained, AI-enabled triage tool can closely replicate GP urgency assessments in the context of same-day appointment requests at the front door of the NHS. The high level of concordance observed between AI and GP ratings demonstrates that AI can serve as a safe and effective adjunct to clinical decision-making. These findings support the safety case for intelligent navigation, a concept of considerable current relevance. The absence of significant under-triage or safety events further supports its potential utility in enhancing patient safety and operational efficiency.

However, successful deployment at scale requires careful attention to key factors, such as the need for robust and continual data collection and performance monitoring to understand the AI tool’s performance. These findings reinforce the need for ongoing human oversight, particularly in edge cases where clinical nuance is paramount.

Despite these challenges, the evidence suggests that AI-enabled triage can enhance access, optimise resource allocation and support sustainable models of primary care especially in overstretched systems like the NHS.

## Supporting information

Appemdix

## Data Availability

The data that support the findings of this study are available from the corresponding author, AEO, upon reasonable request

## Declarations

### Patient and Public Involvement

No patients/patients were involved

## Acknowledgements

The authors thank the staff from Wealden Ridge Medical Partnership for their generous contributions to this study.

## Author Contributors

All authors provided substantial contributions to the conception (AEO, ERS, SA, MA, KL, AP, RR), design (AEO, ERS, SA, MA, AP, RR), acquisition (AEO, ERS SA, MA, KL, AP, RR) and interpretation (SA, MA, ERS, AEO, KL, AP, RR) of study data and approved the final version of the paper. AEO took the lead in planning the study with support from co-authors. SA and ERS carried out the data analysis with support from AEO and MA. AEO is the guarantor.

## Competing interests

KL, AP and HG are employees of Visiba. RR is an employee of WRMP. The other authors did not declare any interests.

**Twitter:** @austenelosta @ImperialSCARU @Sami_Altalib

