## Supplementary material for "Benchmarking Artificial Intelligence vs General Practitioners Decision-Making in Same-Day Appointments Triage: A Mixed-Methods Study in UK Primary Care": Appemdix

### APPENDIX

#### Supplementary Table 1: Modified Manchester Triage System

| Score | Maximum waiting time for consultation | Urgency category |
|---|---|---|
| 1 | 0 minutes | Emergency |
| 2 | 10 minutes | Emergency |
| 3 | 60 minutes | Emergency |
| 4 | 4 hours | Within 24 hours |
| 5 | 1 day | Within 24 hours |
| 6 | 1 week | Non-urgent |
| 7 | 1 month | Non-urgent |
| 8 | Self-care | Non-urgent |

### Supplementary Table 2: Semi structured interview guide

| | |
|---|---|
| **Background information** | • Could you tell me a little bit about yourself (years of experience, role, region/area where you trained/worked)? |
| **Motivations to start/use** | • How did you decide to start using Visiba?<br>• Why Visiba? Other options? |
| **Experience using the triage system** | • What do you think of the Visiba tool/AI-enabled triage? Could you tell me a bit about your experience using the system? Could you please briefly describe the process?<br>• How did you first hear about the AI-enabled triage? What motivated you to use the system?<br>• How long have you been using it? Approximately how many triage cases have you seen so far?<br>• Do you plan to continue using it in the future?<br>• Had you used a similar system before? If so, please tell us more, in particular what similar system did you use, was the alternative better/worse and why? In which context? |
| **Perceived impact** | • Was there anything you found particularly useful or interesting? Any specific benefits of using it? On a scale of 1 - 10, how would you rate the usefulness of the triage tool in helping you triage appointment urgency? Why do you give it this rating?<br>• Was there anything about this triage system that you didn't like so much? Is there something you think should be changed? If so, what?<br>• How do you think the system impacts your clinical work (triage, diagnostic, etc)? In particular, how does having a supporting list of differential diagnosis affect your clinical confidence?<br>• How do you think it impacts your workload? Do you feel the use of the triage tool makes your workload more or less manageable, and why?<br>• How does it impact the appropriateness of same-day appointments?<br>• How the introduction of the tool has changed ways of working- e.g. required extra task of processing triage requests for duty clinician- how they need time allocated to do this but reduces extra slots being booked into clinics. Or how receptionists now need to spend some time doing proxy flows with patients?<br>• Any other effects you could think of? |
| **Confidence & accessibility** | • How confident do you feel about using the system?<br>• How much training was required to start using the system/how intuitive is it?<br>• Do you think it provides accurate triage? What do you think might affect the system's accuracy? |

| | |
|---|---|
| | • Are your clinical decisions supported by using the tool? How?<br>• Do you think it is safe to use? Are there any risks you could think of which could rise from its use?<br>• In particular, how does using this tool affect your tolerance of clinical risk (if any effect)? And what factors allow you to close a case without requiring to contact a patient? |
| **Acceptability & accessibility** | • Do you think it is acceptable and accessible to all patients? What do you think might affect its acceptability? Its accessibility?<br>• What specific group of patients do you feel are affected the most positively/negatively by the 1) system tool and the 2) new way of working? Eg: age or abilities of patients?<br>• Do you think it would be preferable to process all same-day appointment requests via Visiba (rather than just using it when the appointments are full)? Or do you think it would be preferable for all appointment requests to be processed via Visiba?<br>• How would you manage the fact that not everyone can use the service- what workarounds or pathways would you put in place to ensure everyone still has access to care? |
| **Recommendations / closure** | • Would you recommend Visiba tool/AI-enabled triage to other GP practices? And why? Could you specify setting, acuity, specialisms, etc.?<br>• What do you think is the potential for Visiba in WRMP, i.e. how would you expand Visiba in WRMP to maximise impact if you feel it had a positive one?<br>• Do you have any recommendations to improve the system? Is there anything you would change? What extra feature or tool would make it more useful to you?<br>• What else do you think should change/be improved to improve patient triage, including the appropriateness of same-day appointments?<br>• How do you generally feel about AI generally and in triage and how these have been impacted since using Visiba? Has using the tool change your views on AI clinical tools? why?<br>• Is there anything we missed or you would like to add? |

#### Supplementary Table 3: Distribution of Patient Feedback Scores Following Use of the AI-Enabled Triage Tool (N=649)

| Feedback score (N= 649) | Rating | Frequency | Percentage |
|---|---|---|---|
| 1 | Very poor | 30 | 10.7 |
| 2 | Poor | 22 | 7.9 |
| 3 | Good | 46 | 16.4 |
| 4 | Very good | 54 | 19.3 |
| 5 | Excellent | 128 | 45.7 |
| Missing | - | 369 | - |

### STROBE Checklist

| Item | Recommendation | Response |
|---|---|---|
| 1 | Indicate the study's design with a commonly used term in the title or the abstract | Title and abstract describe the study as a 'mixed-methods study in UK primary care', with explanation of quantitative and qualitative components. |
| 2 | Explain the scientific background and rationale for the investigation being reported | Background provides rationale for evaluating AI triage tools compared with GP decision-making in same-day appointment triage. |
| 3 | State specific objectives, including any prespecified hypotheses | Aims clearly stated: assess agreement between AI and GP triage, evaluate safety, usability and perceptions. |
| 4 | Present key elements of study design early in the paper | Methods section specifies explanatory sequential mixed-methods design involving cross-sectional analysis and GP interviews. |
| 5 | Describe the setting, locations, and relevant dates, including periods of recruitment, exposure, follow-up, and data collection | Study conducted across four NHS practices in East Sussex between Jan–Jun 2024, with interviews in Aug 2024. |
| 6 | Give the eligibility criteria, and the sources and methods of selection of participants | All users of the AI triage tool in the 6-month period included. No exclusion criteria applied. Eight GPs recruited for interviews. |
| 7 | Clearly define all outcomes, exposures, predictors, potential confounders, and effect modifiers | Primary outcome: concordance between AI and GP triage scores. Secondary outcomes: satisfaction, perceptions of accuracy/safety. Confounders: age, comorbidities, digital access. |
| 8 | For each variable of interest, give sources of data and details of methods of assessment (measurement) | Urgency ratings from AI and GP recorded. Demographic data extracted from EHR. Interviews used semi-structured guide (Appendix 1). |
| 9 | Describe any efforts to address potential sources of bias | Bias mitigation via random sampling for GP review; clinician oversight of AI scores; limitations in discussion acknowledge bias from proxy users and incomplete data. |
| 10 | Explain how the study size was arrived at | Sample size calculation using OpenEpi with CI 99% & population size 2,671. Target: 532, actual: 649. |
| 11 | Explain how quantitative variables were handled in the analyses | Urgency scores analysed as ordinal. Age grouped. Logistic regression used for satisfaction. |
| 12 | Describe all statistical methods, including those used to control for confounding | Spearman's ρ, Cohen's kappa, ordinal logistic regression performed. Age and gender used in adjusted models. STATA v17 used. |
| 13 | Report numbers of individuals at each stage of study | Flow of SDA requests shown in Figure 1. Table 1 provides demographic breakdown. Feedback response N=280. |
| 14 | Give characteristics of study participants | Detailed in Table 1 (age, gender, ethnicity). Additional qualitative participant profile given. |

| | | |
|---|---|---|
| **15** | Report numbers of outcome events or summary measures | Urgency ratings compared in Table 2. Agreement in Table 3. Confusion matrix in Table 4. Satisfaction in Table 5. |
| **16** | Give unadjusted and adjusted estimates and their precision | Ordinal regression results in Table 5 with aOR, 95% CI & p-values for satisfaction predictors. |
| **17** | Report other analyses done | Thematic analysis of GP interviews conducted using Framework Method. Themes: accuracy, safety, usability. |
| **18** | Summarize key results with reference to study objectives | Findings summarised in Abstract & Discussion: AI vs GP agreement substantial; tool conservative & safe; older patients less satisfied. |
| **19** | Discuss limitations of the study | Discussed comprehensively: limited generalisability, proxy bias, lack of EHR access, partial feedback response. |
| **20** | Give a cautious overall interpretation of results | Conclusions balanced: AI triage safe & scalable but requires oversight, EHR integration & digital inclusion strategies. |
| **21** | Discuss the generalisability (external validity) of the study results | Acknowledged: results may not apply beyond East Sussex NHS settings; future replication needed in diverse contexts. |

### COREQ Checklist

| Domain | Item | Guide Question/Description | Response |
|---|---|---|---|
| **Research team and reflexivity** | 1. Interviewer/facilitator | Which author conducted the interview or focus group? | Interviews were conducted by AEO and ERS. |
| **Research team and reflexivity** | 2. Credentials | What were the researcher's credentials? | AEO holds MSc, MPA, PhD; ERS holds MA. |
| **Research team and reflexivity** | 3. Occupation | What was their occupation at the time of the study? | AEO and ERS were affiliated with Imperial College London as researchers. |
| **Research team and reflexivity** | 4. Gender | Was the researcher male or female? | AEO is male; ERS is female. |
| **Research team and reflexivity** | 5. Experience and training | What experience or training did the researcher have? | Both researchers had training in qualitative methods and prior experience conducting interviews in healthcare research. |
| **Study design** | 6. Relationship established | Was a relationship established prior to study commencement? | No prior relationship with participants beyond professional interaction within the study setting. |
| **Study design** | 7. Participant knowledge of the interviewer | What did the participants know about the researcher? | Participants were provided with a participant information sheet describing the study purpose and roles of the researchers. |
| **Study design** | 8. Interviewer characteristics | What characteristics were reported about the interviewer/facilitator? | Researchers disclosed their academic roles and interest in evaluating AI-enabled triage tools. |
| **Study design** | 9. Methodological orientation | What methodological orientation was stated to underpin the study? | Thematic analysis using the Framework Method was employed. |
| **Study design** | 10. Sampling | How were participants selected? | Convenience sampling of GPs who had used the AI tool for 6–8 months. |
| **Study design** | 11. Method of approach | How were participants approached? | Via email invitation and provision of participant information sheet. |
| **Study design** | 12. Sample size | How many participants were in the study? | Eight GPs participated. |
| **Study design** | 13. Non-participation | How many people refused or dropped out? Reasons? | None; all approached GPs agreed to participate. |

| | | | |
|---|---|---|---|
| **Study design** | 14. Setting of data collection | Where was the data collected? | Interviews conducted online via MS Teams. |
| **Study design** | 15. Presence of non-participants | Was anyone else present besides the participants and researchers? | No, only researchers and participants were present. |
| **Study design** | 16. Description of sample | What are the important characteristics of the sample? | All participants were practising GPs, 6 male and 2 female, from East Sussex NHS practices. |
| **Data collection** | 17. Interview guide | Were questions, prompts, guides provided? Was it pilot tested? | A semi-structured interview guide was used and reviewed internally; not pilot tested. |
| **Data collection** | 18. Repeat interviews | Were repeat interviews carried out? | No repeat interviews were conducted. |
| **Data collection** | 19. Audio/visual recording | Did the research use audio or visual recording to collect the data? | Yes, all interviews were audio-recorded. |
| **Data collection** | 20. Field notes | Were field notes made during and/or after the interview or focus group? | Field notes were taken during the interviews. |
| **Data collection** | 21. Duration | What was the duration of the interviews or focus groups? | Interviews lasted 45 to 55 minutes. |
| **Data collection** | 22. Data saturation | Was data saturation discussed? | Data saturation was not formally discussed but thematic convergence was observed. |
| **Data collection** | 23. Transcripts returned | Were transcripts returned to participants for comment and/or correction? | No, transcripts were not returned to participants. |
| **Data analysis and findings** | 24. Number of data coders | How many data coders coded the data? | Two researchers (AEO and ERS) coded the data. |
| **Data analysis and findings** | 25. Description of the coding tree | Did authors provide a description of the coding tree? | Codes and themes were organised using the Framework Method but not all coding trees were presented. |
| **Data analysis and findings** | 26. Derivation of themes | Were themes identified in advance or derived from the data? | Themes were derived inductively from the data. |
| **Data analysis and findings** | 27. Software | What software, if applicable, was used to manage the data? | Data was manually coded using Microsoft Word and Excel. |
| **Data analysis** | 28. Participant checking | Did participants provide feedback on the findings? | No, participant checking was not performed. |

| | | | |
|---|---|---|---|
| **and findings** | | | |
| **Data analysis and findings** | 29. Quotations presented | Were participant quotations presented to illustrate the themes/findings? | Yes, quotations were included to illustrate key themes. |
| **Data analysis and findings** | 30. Data and findings consistency | Was there consistency between the data presented and the findings? | Yes, findings were clearly supported by interview quotations. |
| **Data analysis and findings** | 31. Clarity of major themes | Were major themes clearly presented in the findings? | Yes, major themes such as safety, accuracy, usability were clearly described. |
| **Data analysis and findings** | 32. Clarity of minor themes | Is there a description of diverse cases or discussion of minor themes? | Yes, minor themes like digital exclusion and proxy use were described. |
